# The accuracy of Japanese administrative data in identifying acute exacerbation of idiopathic pulmonary fibrosis

**DOI:** 10.1101/2021.07.04.21259968

**Authors:** Keisuke Anan, Yuki Kataoka, Kazuya Ichikado, Kodai Kawamura, Takeshi Johkoh, Kiminori Fujimoto, Kazunori Tobino, Ryo Tachikawa, Hiroyuki Ito, Takahito Nakamura, Tomoo Kishaba, Minoru Inomata, Yosuke Yamamoto

## Abstract

**Background:** This study aimed to develop criteria for identifying patients with acute exacerbation of idiopathic pulmonary fibrosis (AE-IPF) from Japanese administrative data and validate the pre-existing criteria.

**Methods:** This retrospective, multi-center validation study was conducted at eight institutes in Japan to verify the diagnostic accuracy of the disease name for AE-IPF. We used the Japanese Diagnosis Procedure Combination data to identify patients with a disease name that could meet the diagnostic criteria for AE-IPF, who were admitted to the eight institutes from January 2016 to February 2019. As a reference standard, two respiratory physicians performed a chart review to determine whether the patients had a disease that met the diagnostic criteria for AE-IPF. Furthermore, two radiologists interpreted the chest computed tomography findings of cases considered AE-IPF and confirmed the diagnosis. We calculated the positive predictive value (PPV) for each disease name and its combination.

**Results:** We included 830 patients; among them, 216 were diagnosed with AE-IPF through the chart review. We combined the groups of disease names and yielded two criteria: the criteria with the highest PPV (0.72 [95% confidence interval 0.62 to 0.81]) and that with a slightly less PPV (0.61 [0.53 to 0.68]) but more true positives. Pre-existing criteria showed a PPV of 0.40 (0.31 to 0.49).

**Conclusion:** The criteria derived in this study for identifying AE-IPF from Japanese administrative data show a fair PPV. Although these criteria should be carefully interpreted according to the target population, our findings could be utilized in future database studies on AE-IPF.

## Background

Idiopathic pulmonary fibrosis (IPF) is a rare disease with a poor prognosis that is characterized by chronic progressive lung fibrosis. Although IPF is rare, it has a worse prognosis than numerous cancer types.[1] Moreover, hospitalization and mortality rates attributable to IPF have increased over recent years, which is suggestive of an increasing burden of this disease on healthcare services worldwide.[2-4] During the disease course, acute exacerbation (AE) may occur at an annual frequency of approximately 5–15%; further, the mortality rate is reported to be approximately 50–60%.[5, 6] Few randomized controlled trials (RCTs) with limited external validity have investigated AE-IPF.[7, 8] Therefore, there is a need for large observational studies on a wider patient population using real-world data to complement RCTs.[9]

However, there have been rare seen large-scale database studies on AE-IPF. This could be attributed to the difficulty in identifying cases. In large-scale database research, case identification is essential, with validation studies being crucial for this purpose. The REporting of studies Conducted using Observational Routinely collected health Data (RECORD) statement mentions that “Any validation studies of the codes or algorithms used to select the population should be referenced.”[10] Regarding AE-IPF, one validation study has been performed in the US[11]; however, the positive predictive value (PPV) was low (0.62 [95% confidence interval [CI] 0.53 to 0.70]). Additionally, it remains unclear whether the data can be applied in other countries.

This study aimed to develop criteria for identifying patients with AE-IPF, as well as validate the criteria in the previous study, using multi-center chart data in Japan.

## Methods

We performed and reported this study in accordance with the reporting guidelines for assessing the quality of validation studies of health administrative data (Table S1).[12]

### Settings and patients

This retrospective, multicenter, validation study was conducted to verify the diagnostic accuracy of the disease name for AE-IPF. We performed study in eight Japanese tertiary hospitals: Saiseikai Kumamoto Hospital, Hyogo Prefectural Amagasaki General Medical Center, Iizuka Hospital, Kobe City Medical Center General Hospital, Kameda Medical Center, Hoshigaoka Medical Center, Okinawa Chubu Hospital, and the Japanese Red Cross Medical Center.

We included patients aged ≥ 40 years with a disease name that could meet the diagnostic criteria for AE-IPF (Table 1), who were admitted to the aforementioned eight institutes from January 2016 to February 2019. These criteria were developed in advance by two respiratory specialists. We excluded patients with disease names for secondary interstitial pneumonia, including collagen vascular disease-associated interstitial lung disease and chronic and hypersensitivity pneumonitis, as well as those for malignant neoplasms (Table 2). We did not exclude patients with disease names for malignant neoplasms with the suffixes “post-operation” or “post-radiotherapy”. We ignored all “suspected” disease names.

**Table 1.**
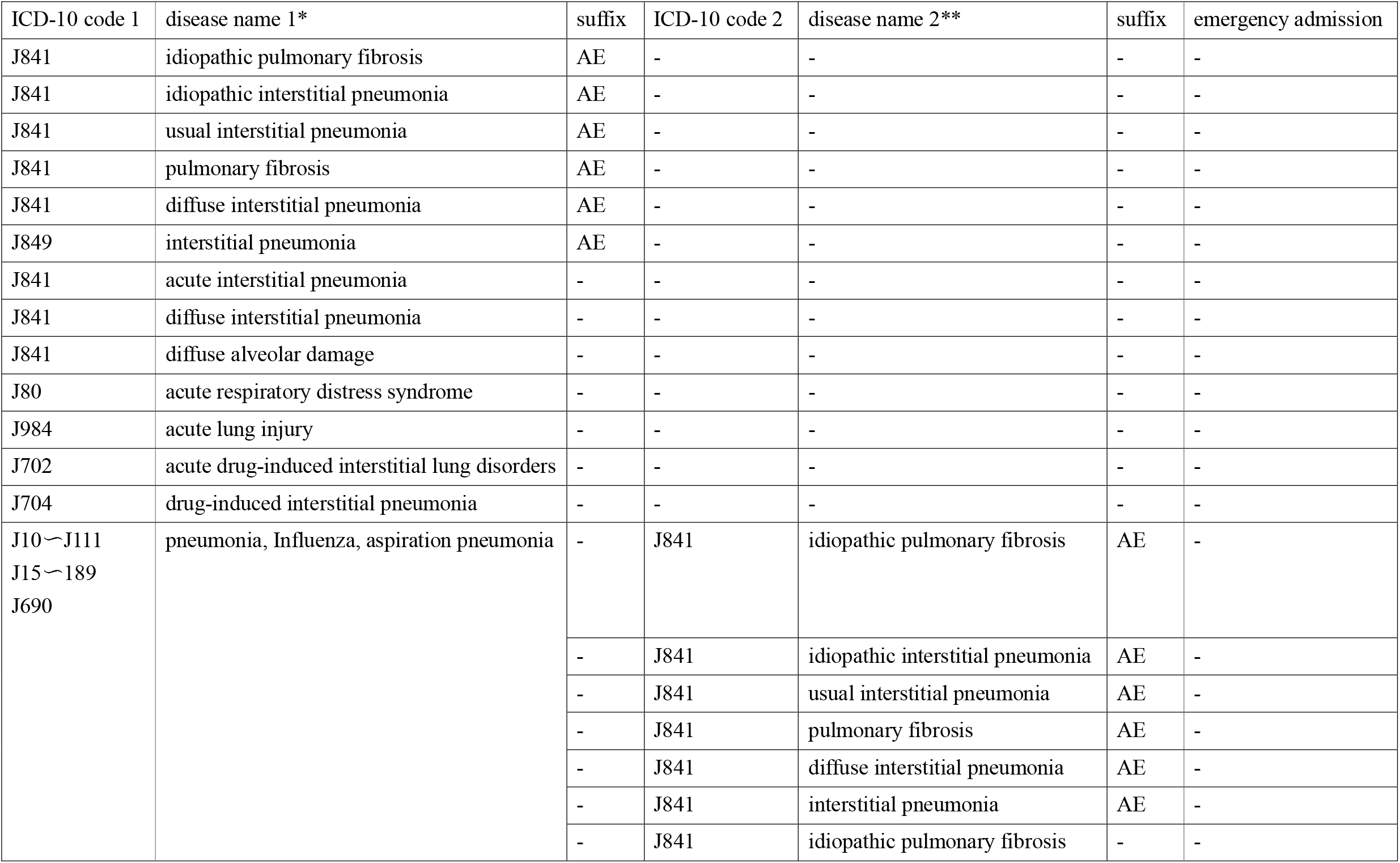
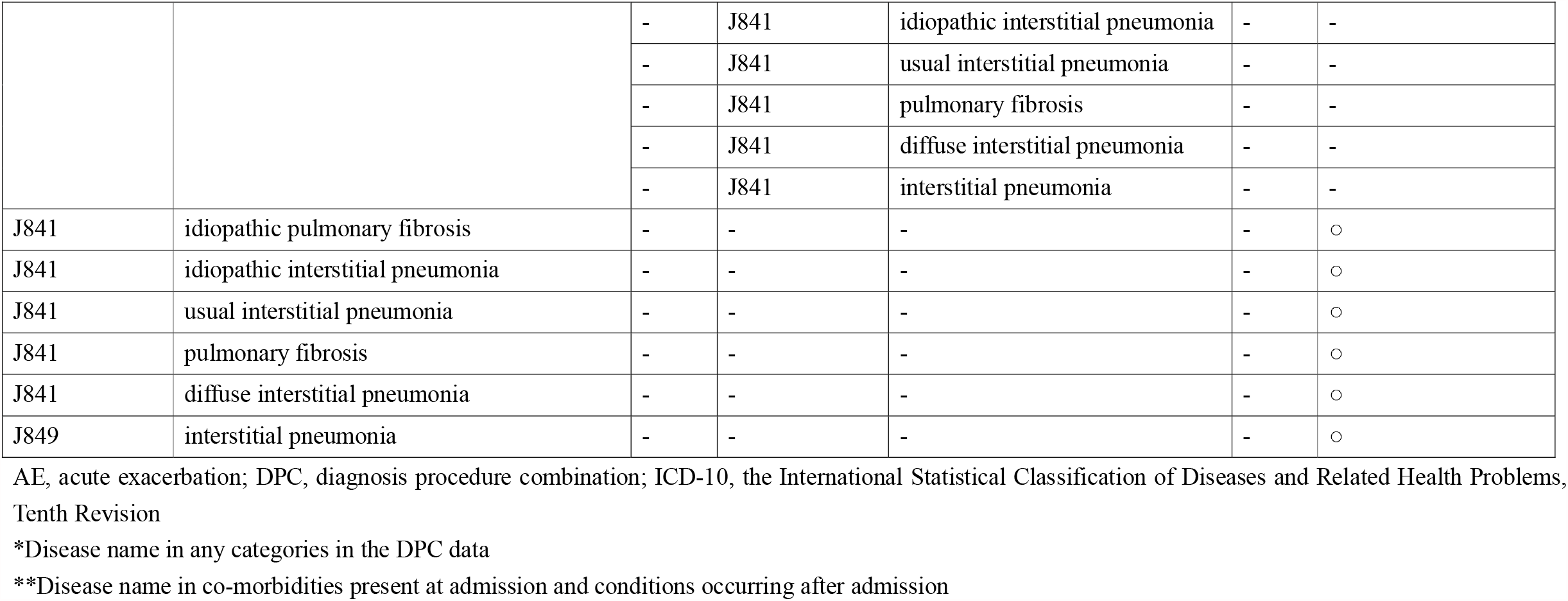
Disease names adopted as inclusion criteria

**Table 2.** Disease names adopted as exclusion criteria

| ICD-10 code | disease name* |
| --- | --- |
| C00～C96 | malignant neoplasms** |
| D860 | sarcoidosis of lung |
| J60 | coal worker's pneumoconiosis |
| J61 | asbestosis |
| J64 | pneumoconiosis |
| J67 | hypersensitivity pneumonitis due to organic dust |
| J990・M0510 | rheumatoid arthritis-associated interstitial lung disease |
| J991・M351 | collagen vascular disease-associated interstitial lung disease |
| J991・M330 | juvenile dermatomyositis-associated interstitial lung disease |
| J991・M321 | systemic lupus erythematosus-associated interstitial lung disease |
| J991・M332 | polymyositis-associated interstitial lung disease |
| J991・M331 | dermatomyositis-associated interstitial lung disease |
| J991・M313 | lung involvement in granulomatosis with polyangiitis |
| M348 | lung involvement in systemic sclerosis |

\* Disease name in any category in the DPC data
\*\*not exclude if they have attached suffix of “post-operation” or “post-radiotherapy”.

Regarding disease names, we extracted data from the diagnosis procedure combination (DPC), which was launched at Japanese acute-care hospitals across the country in 2003.[13, 14] The following data were extracted from the DPC: some clinical information (e.g. age, gender, etc.), admission route (e.g. scheduled or emergency admission), diagnoses, surgeries and procedures, medications, outcomes, etc. The patients were recorded with diagnostic names using the International Statistical Classification of Diseases and Related Health Problems, Tenth Revision (ICD-10) codes. The diagnoses included the following six categories: main diagnosis, diagnosis that triggered hospitalization, diagnosis using the most medical resources, diagnosis using the second most medical resources, comorbidities present at admission, and conditions occurring after admission. One diagnostic name is coded for each main diagnosis, diagnosis that triggered hospitalization, diagnosis using the most medical resources, and diagnosis using the second most medical resources. Meanwhile, up to 10 diagnostic names can be coded for comorbidities present at admission and conditions occurring after admission. Furthermore, in the Japanese DPC data, modifiers can be added to the disease name in ICD-10 (e.g., regions such as left and right, recurrence, acute exacerbation, suspected, etc.).

This study was approved by the Ethics Committee of leading institute of this study (Kyoto University Graduate School and Faculty of Medicine) (Approved Number: R2071) and other committed institutes. The requirement for written informed consent from the participants was waived because of the retrospective design and use of anonymized data.

### Reference standard

We confirmed the AE-IPF diagnosis using the following two steps:

1. At least two respiratory physicians at each institute reviewed the medical records and chest computed tomography (CT) findings on admission. Subsequently, they determined whether the patients met the following diagnostic criteria: an official ATS/ERS/JRS/ALAT clinical practice guideline for IPF diagnosis [15] and the international working group diagnostic criteria for AE-IPF.[16] An IPF diagnosis was determined if UIP or probable UIP pattern was observed on HRCT findings, even without surgical lung biopsy (SLB). If SLB was performed, IPF was considered based on specific combinations of HRCT and histopathological patterns following the ATS/ERS/JRS/ALAT clinical practice guidelines.[15] Discrepancies were resolved through discussion or consultation with a third respiratory physician, as appropriate.
2. Two expert chest radiologists interpreted chest CT findings of cases considered to be AE-IPF in 1) and determined whether the patients met the aforementioned two diagnostic criteria. If both radiologists considered that a case was not AE-IPF, it was determined as not AE-IPF. On the other hand, if a case was considered as not as AE-IPF by only one radiologist, the case was determined as AE-IPF.

Multidisciplinary discussion (MDD), which is held for diagnostic decision-making regarding patients with interstitial lung disease who are clinically suspected to have IPF, has been described in the official ATS/ERS/JRS/ALAT clinical practice guideline.[15] However, MDD is a conditional recommendation that is not widely held in Japanese hospitals. Therefore, we decided not to consider MDD as a diagnostic requirement.

### Developing criteria

First, we assessed the diagnostic accuracy of each disease name in the inclusion criteria, with disease names showing high diagnostic accuracy being adopted without modification. Next, we explored combinations of disease names with high diagnostic accuracy for disease names with a high number of true positives. We sought to create two criteria: 1) the criteria with the highest PPV (specificity-maximizing criteria) and 2) criteria with slightly less PPV but more true positives (balanced criteria). Based on previous studies, [11, 17, 18] we excluded disease names of secondary interstitial lung diseases (ILDs) and included those of acute respiratory failure (ARF) (Table 3). Further, we excluded disease names with pneumonia, influenza, and aspiration pneumonia and addition of that of IPF.

**Table 3.** ICD-10 codes for interstitial lung diseases (ILDs) other than idiopathic pulmonary fibrosis (IPF) and acute respiratory failure.

| ILDs other than IPF |  |
| --- | --- |
| ICD-10 code | disease name |
| C966 | Langerhans-cell histiocytosis* |
| D869 | Sarcoidosis |
| E756 | Lipidoses |
| J998 • E831 | Idiopathic pulmonary hemosiderosis |
| E85 | Amyloidosis |
| J60 | coal worker's pneumoconiosis* |
| J61 | asbestosis* |
| J62 | silicosis or talcosis |
| J63 | berylliosis and other inorganic dusts |
| J64 | unspecified pneumoconiosis* |
| J67 | hypersensitivity pneumonitis* |
| J82 | Pulmonary eosinophilia, not elsewhere classified |
| J82 | Extrinsic allergic alveolitis |
| J684 | Chronic respiratory conditions due to chemicals, gases, fumes, and vapors |
| J701 | Chronic and other pulmonary manifestations due to radiation |
| J708 | Respiratory conditions due to other specified external agents |
| J840 | Alveolar and parieto-alveolar conditions |
| J841 | Postinflammatory pulmonary fibrosis |
| J99 | Respiratory disorders in diseases classified elsewhere |
| K50 | Regional enteritis |
| M05, M06 | Rheumatoid arthritis |
| M310 | Goodpasture' s syndrome |
| M313 | Wegener' s granulomatosis |
| M317 | Microscopic polyangiitis |
| M318 | Other specified necrotizing vasculopathies |
| M321, M329 | Systemic lupus erythematosus |
| M330, M331, M339 | Dermatomyositis |
| M332 | Polymyositis |
| M340, M341, M349 | Systemic sclerosis |
| M348 | Lung involvement in systemic sclerosis* |
| M350 | Sjogren' s disease |
| M351 | other overlap syndrome |
| M45 | Ankylosing spondylitis |
| Q850 | Neurofibromatosis |
| Q851 | Tuberous sclerosis |
| Acute respiratory failure |  |
| ICD-10 code | disease name |
| J960 | acute respiratory failure |

\*We already excluded these disease names as exclusion criteria

### External validation of the criteria of a previous study

To compare the PPV with the aforementioned newly developed criteria, we externally validated the criteria of a previous study [11] (pre-existing criteria) using DPC data extracted from the aforementioned eight hospitals. The inclusion criteria were as follows: 1) age ≥ 50 years; 2) having at least one claim to a specific diagnostic code for IPF (either ICD10 code J84.9, J84.10, J84.111, J84.112, or ICD-9 correspond); 3) having acute respiratory symptoms or ARF (J84.114, J84.9, J96.00-02, J96.20-22, J96.91-92, or ICD-9 correspond) and subsequent hospital admission; and 4) lacking secondary ILDs.[11] In accordance with the Japanese DPC data used in this study, we modified this criteria as follows: 1) age ≥ 50 years; 2) having at least one following disease name (either ICD10 code J84.9 [interstitial pneumonia (IP)] or J84.1 [IPF or idiopathic interstitial pneumonia (IIP) or pulmonary fibrosis] in three diagnosis categories (main diagnosis, diagnosis that triggered hospitalization, diagnosis using the most medical resources); 3) having at least one following disease names (ICD10 code J84.1 [acute interstitial pneumonia] or J96.0 [ARF]) in any diagnosis category, and emergency admission; and 4) lacking secondary ILDs.

### Statistical analysis

Interobserver agreement for the AE-IPF diagnosis between both respiratory specialists was evaluated using kappa statistics. For cases diagnosed as AE-IPF by two respiratory physicians, the interobserver agreement for AE-IPF diagnosis between both radiologists was assessed using the AC_1_-index since most cases were considered to be true positive cases.[19] We calculated the PPV as true positives / (true positives + false positives) for every disease name and their combinations. Further, we calculated the 95% confidence intervals (CIs) for binomial distribution using the exact method. We could not calculate the sensitivity, specificity, and negative predictive values since we did not perform a chart review of patients without target ICD-10 codes. Validation studies for only obtaining a PPV have reported that a sample size of approximately 100 is sufficient.[20] We sought to identify patients from various disease names and search for algorithms that would improve PPV. Therefore, regarding the specificity-maximizing criteria, we identified the final criteria with the largest PPV out of approximately 100 cases. Statistical analyses were performed using STATA/SE version.16.0 (Stata Corp., College Station, TX, USA).

## Results

This study included 822 patients; among them, 212 were diagnosed with AE-IPF by chart review. There was substantial interobserver agreement for AE-IPF diagnosis (kappa = 0.80 [0.75 to 0.85]). Among the patients who were considered to have AE-IPF by both respiratory physicians, none were considered not to have AE-IPF by both radiologists; moreover, there was excellent interobserver agreement between both radiologists (AC_1_-index = 0.99 [0.98 to 1]).

Table 4 shows the PPV and 95% CI for each disease name. Although the disease name of AE-IPF and acute exacerbation of IIP (AE-IIP) had the highest and second-highest PPV, respectively, the number of patients was small. The PPV for all the other disease names was low.

**Table 4.** The positive predictive value (PPV) and 95% confidence intervals (CI) of patients diagnosed with acute exacerbation of idiopathic pulmonary fibrosis within each algorithm

| disease names | total (n) | True Positives | PPV | 95% CI |
| --- | --- | --- | --- | --- |
| IPF + AE | 5 | 4 | 0.8 | 0.28 to 0.99 |
| IIP + AE | 32 | 24 | 0.75 | 0.57 to 0.89 |
| (IIP + AE) - secondary ILDs* | 30 | 23 | 0.77 | 0.58 to 0.90 |
| (IIP + AE) - secondary ILDs + ARF* | 22 | 17 | 0.77 | 0.55 to 0.92 |
| (IIP + AE) - secondary ILDs - infection | 29 | 22 | 0.76 | 0.55 to 0.92 |
| (IIP + AE) - secondary ILDs - infection + ARF | 21 | 16 | 0.76 | 0.56 to 0.90 |
| (IIP + AE) + IPF | 6 | 6 | 1 | 0.54 to 1 |
| UIP + AE | 0 | 0 | - | - |
| pulmonary fibrosis + AE | 2 | 1 | 0.5 | 0.13 to 0.99 |
| diffuse IP + AE | 8 | 1 | 0.13 | 0.003 to 0.53 |
| IP + AE | 105 | 59 | 0.56 | 0.46 to 0.66 |
| (IP + AE) - secondary ILDs | 95 | 57 | 0.6 | 0.49 to 0.70 |
| (IP + AE) - secondary ILDs + ARF | 75 | 50 | 0.67 | 0.55 to 0.77 |
| (IP + AE) - secondary ILDs - infection | 77 | 48 | 0.62 | 0.51 to 0.73 |
| (IP + AE) - secondary ILDs - infection + ARF | 60 | 42 | 0.7 | 0.57 to 0.81 |
| (IP + AE) + IPF | 15 | 9 | 0.6 | 0.32 to 0.84 |
| (IP + AE) + IPF - secondary ILDs - infection | 12 | 8 | 0.67 | 0.35 to 0.90 |
| AIP | 40 | 11 | 0.28 | 0.15 to 0.44 |
| diffuse IP | 86 | 8 | 0.093 | 0.041 to 0.18 |
| DAD | 1 | 0 | 0 | 0 to 0.98 |
| ARDS | 66 | 5 | 0.076 | 0.025 to 0.17 |
| ALI | 2 | 0 | 0 | 0 to 0.84 |
| acute drug-induced interstitial lung disorders | 13 | 1 | 0.077 | 0.002 to 0.36 |
| drug-induced interstitial pneumonia | 47 | 2 | 0.043 | 0.005 to 0.15 |
| Infection + (IPF + AE) | 0 | 0 | - | - |
| Infection + (IIP + AE) | 1 | 0 | 0 | 0 to 0.98 |
| Infection + (UIP + AE) | 0 | 0 | - | - |
| Infection + (pulmonary fibrosis + AE) | 0 | 0 | - | - |
| Infection + (diffuse IP+AE) | 0 | 0 | - | - |
| Infection + (IP + AE) | 11 | 4 | 0.36 | 0.11 to 0.69 |
| Infection + IPF | 15 | 3 | 0.2 | 0.043 to 0.48 |
| Infection + IIP | 15 | 3 | 0.2 | 0.043 to 0.48 |
| Infection + UIP | 0 | 0 | - | - |
| Infection + pulmonary fibrosis | 5 | 0 | 0 | 0 to 0.52 |
| Infection + diffuse IP | 3 | 0 | 0 | 0 to 0.71 |
| Infection + IP | 82 | 11 | 0.13 | 0.069 to 0.23 |
| IPF + emergency admission | 40 | 16 | 0.4 | 0.25 to 0.57 |
| IPF + emergency admission + ARF | 15 | 10 | 0.67 | 0.38 to 0.88 |
| IPF + emergency admission - secondary ILDs - infection + ARF | 12 | 8 | 0.67 | 0.35 to 0.90 |
| IPF + emergency admission + (IP + AE) | 4 | 2 | 0.5 | 0.068 to 0.93 |
| IIP + emergency admission | 152 | 35 | 0.23 | 0.17 to 0.31 |
| IIP + emergency admission + ARF | 47 | 19 | 0.40 | 0.26 to 0.56 |
| IIP + emergency admission - secondary ILDs - infection + ARF | 21 | 7 | 0.33 | 0.15 to 0.57 |
| IIP + emergency admission + (IP + AE) | 3 | 2 | 0.67 | 0.094 to 0.99 |
| UIP + emergency admission | 11 | 4 | 0.36 | 0.11 to 0.69 |
| pulmonary fibrosis + emergency admission | 3 | 0 | 0 | 0 to 0.71 |
| diffuse IP + emergency admission | 53 | 7 | 0.13 | 0.055 to 0.25 |
| IP + emergency admission | 140 | 38 | 0.27 | 0.20 to 0.35 |
| IP + emergency admission + ARF | 78 | 27 | 0.35 | 0.24 to 0.46 |
| IP + emergency admission - secondary ILDs - infection + ARF | 45 | 14 | 0.31 | 0.18 to 0.47 |
| final algorithm |  |  |  |  |
| Specificity maximizing criterion |  |  |  |  |
| (IPF + AE) OR ((IIP + AE) - secondary ILDs - infection) OR ((IIP + AE) + IPF) OR ((IP + AE) - secondary ILDs - infection + ARF) | 94 | 68 | 0.72 | 0.62 to 0.81 |
| Balanced algorithm |  |  |  |  |
| (IPF + AE) OR (IIP + AE) OR ((IP + AE) - secondary ILDs + ARF) OR (IPF + emergency admission + (ARF OR (IP + AE))) OR (IIP + emergency admission + (ARF OR (IP + AE))) | 168 | 102 | 0.61 | 0.53 to 0.68 |
| Pre-existing criterion |  |  |  |  |
| ((IPF OR IIP OR pulmonary fibrosis OR IP) + (AIP OR ARF) + emergency admission - secondary ILDs) | 126 | 50 | 0.40 | 0.31 to 0.49 |

AE, acute exacerbation; AIP, acute interstitial pneumonia; ALI, acute lung injury; ARDS, acute respiratory distress syndrome; ARF, acute respiratory failure; DAD, diffuse alveolar damage; IIP, idiopathic interstitial pneumonia; ILDs, interstitial lung diseases; IP, interstitial pneumonia; IPF, idiopathic pulmonary fibrosis; UIP, usual interstitial pneumonia
\*disease names of secondary ILDs and ARF are described in Table 3

### Specificity-maximizing criteria

We explored algorithms that would result in higher PPV for the five disease names (AE-IIP, acute exacerbation of interstitial pneumonia, AE-IP, IPF + emergency admission, IIP + emergency admission, and IP + emergency admission) and had more true positive patients. We attempted to exclude disease names related to secondary ILDs and infection, and add to disease names for ARF and IPF. Consequently, we observed an increased PPV for several combinations of disease names.

Finally, we combined the groups of disease names with a higher PPV and yielded a criteria (AE-IPF OR (AE-IIP - secondary ILDs - infection) OR (AE-IIP + IPF) OR (AE-IP - secondary ILDs - infection + ARF) with a PPV of 0.72 (95% CI 0.62 to 0.81) (Table 4).

### Balanced criteria

Using the five disease names, we searched for a criteria that would include more true-positive patients even with a slightly inferior PPV. Finally, we created a criteria (AE-IPF OR AE-IIP OR [AE-IP - secondary ILDs + ARF] OR [IPF + emergency admission + [ARF OR AE-IP] OR [IIP + emergency admission + (ARF OR AE-IP)] with a PPV of 0.61 (95% CI 0.53 to 0.68) (Table 4).

### Pre-existing criteria

The validation results for the pre-existing criteria (IPF OR IIP OR pulmonary fibrosis OR IP) + (AIP OR ARF) + emergency admissions - secondary ILDs) showed a PPV of 0.40 (95% CI 0.31 to 0.49) (Table 4).

## Discussion

Using Japanese administrative data, we created criteria for identifying patients with AE-IPF and validated the pre-existing criteria. It was difficult to accurately identify true positive AE-IPF cases only based on a single disease name. Consequently, we excluded or added various other disease names and emergency admissions, which improved the AE-IPF diagnosis accuracy. The pre-existing criteria had a very low PPV. The PPV for the final criteria were as follows: specificity-maximizing criteria, 0.72 (0.62 to 0.81); balanced criteria, 0.61 (0.53 to 0.68); and pre-existing criteria, 0.40 (0.31 to 0.49).

The diagnostic accuracy of the specificity-maximizing and balanced criteria showed a fair PPV. Currently, the only validation study on AE-IPF derived a criteria with a PPV of 0.62 (95% CI 0.53 to 0.70),[11] which was lower and comparable to the PPV of the specificity-maximizing and balanced criteria, respectively. Most disease name combinations in the specificity-maximizing criteria used the suffix acute exacerbation, which is unique to the DPC data and may have attributed to the higher PPV compared with that in the previous study. Meanwhile, the validation results for the pre-existing criteria revealed a very low PPV (0.40 [95% CI 0.31 to 0.49]). This indicates that it may be better to use the specificity-maximizing or balanced criteria rather than the pre-existing criteria in the Japanese DPC data.

Meanwhile, our findings are suggestive of the difficulty of AE-IPF diagnosis only based on administrative data. As shown in Table 4, there were only four true-positive patients with a disease name of AE-IPF. IPF is a rare disease; moreover, its diagnosis is heavily dependent on high-resolution CT (HRCT). Additionally, IPF diagnosis is sometimes difficult since it requires the exclusion of all other diseases that cause chronic fibrosis of the lungs. Furthermore, in case of acute respiratory worsening in fibrotic ILDs, [21] it can be challenging for clinicians to distinguish between AE and others. Therefore, it is very difficult to accurately diagnose AE-IPF; moreover, many true positive patients could have been included among those with disease names of IIP or IP as well as IPF with or without AE. To exclude secondary ILDs, we referred to the ICD-9 and 10 codes used in previous IPF database studies.[11, 17, 18] For AE, we referred to a previous study and added a code for ARF.[11] Combining these codes with IIP and IP increased the PPV; therefore, these codes may be useful for excluding secondary ILDs and including AEs. For diseases that are rare and difficult to diagnose, a PPV of approximately ≥ 70% is considered reasonable [22, 23]. Therefore, given the aforementioned difficulties in diagnosing AE-IPF, the PPVs obtained in our study could be considered reasonable.

### Clinical implications

We believe our findings regarding the criteria for identifying patients with AE-IPF can contribute to database research in Japan. Meanwhile, using the specificity-maximizing criteria, only about 1/3 of the cases were true positives. To detect more patients with AE-IPF, it may be better to use the balanced criteria even though it has a lower PPV. Additionally, the specificity-maximizing and balanced criteria are both insufficient for selecting patients with AE-IPF patients without omission. It should be recognized that some patients with disease names, including acute respiratory distress syndrome or drug-induced interstitial pneumonia, may meet the criteria for AE-IPF. Furthermore, the institutions involved in this study were tertiary hospitals that treat numerous patients with AE-IPF. Although DPC data are collected from numerous acute care hospitals, there were not all proficient in treating patients with AE-IPF; thus, our findings may not be generalizable as representative data for Japan.

### Research implication

We created criteria for defining AE-IPF with a fair PPV, however, it was not high. Although the present study and a previous validation study were conducted only using disease names and hospitalization information, an algorithm with a higher PPV can be employed by adding other information, including blood test and prescription data. Additionally, using PPV alone cannot allow for a database study that covers the entire AE-IPF population. Therefore, there is a need for studies calculating the sensitivity and specificity using methods, including random sampling or “all possible cases” sampling.[24]

### Strength and Limitations

Our study has several strengths. First, this is the first validation study on AE-IPF using Japanese DPC data. Second, this was a multi-center study with a sufficient sample size. Third, AE-IPF diagnosis was more reliable because of the interpretation by two chest radiologists in addition to at least two pulmonologists.

This study had several limitations. First, since this study was conducted at tertiary emergency hospitals with respiratory specialists, it remains unclear whether it can be adapted to the entire DPC data in Japan. Therefore, our findings should be externally validated in other facilities in Japan. Second, this was a validation study of the Japanese DPC data, which is unique to Japan and does not apply to other countries. Finally, we cannot eliminate the possibility of misclassification of the IPF diagnosis (e.g., hypersensitivity pneumonitis and collagen vascular disease) or AE. However, we believe that this was reduced by the interpretation of the chest CT findings by two expert chest radiologists.

## Conclusion

In conclusion, this study derived criteria for identifying AE-IPF from Japanese administrative data with a fair PPV. When studying AE-IPF in Japanese administrative data using the specificity-maximizing or balanced criteria, researchers should carefully interpret the target population.

## Data Availability

The data that support the findings of this study are available from the corresponding author upon reasonable request.

## Acknowledgments

We are grateful to Asuka Iwashita (Saiseikai Kumamoto Hospital), Hiromi Ide (Iizuka Hospital), Keiko Sakuragawa (Kobe City Medical Center General Hospital), Kyoko Wasai (Hyogo Prefectural Amagasaki General Medical Center), Hiroshi Harada (Kameda Medical Center), Megumi Kunita (Hoshigaoka Medical Center), Daiki Chinen (Okinawa Chubu Hospital), and Misao Ebihara (Japanese Red Cross Medical Center) for retrieving data. We would like to thank Editage (www.editage.com) for English language editing.

---

**Table S1.** Checklist of reporting criteria for studies validating health administrative data algorithms.

|  | YES | NO | UNCERTAIN | NOT APPLICABLE |
| --- | --- | --- | --- | --- |
| **TITLE, KEYWORDS, ABSTRACT** |  |  |  |  |
| Identify article as study of assessing diagnostic accuracy | ✓ ▪ |  |  |  |
| Identify article as study of administrative data | ✓ ▪ |  |  |  |
| **INTRODUCTION:** |  |  |  |  |
| State disease identification & validation one of goals of study | ✓ ▪ |  |  |  |
| **METHODS:** |  |  |  |  |
| *Participants in validation cohort:* |  |  |  |  |
| Describe validation cohort (Cohort of patients to which reference standard was applied) | ✓ ▪ |  |  |  |
| · Age |  | ✓ ▪ |  |  |
| · Disease | ✓ ▪ |  |  |  |
| · Severity |  |  |  | ✓ ▪ |
| · Location/Jurisdiction | ✓ ▪ |  |  |  |
| Describe recruitment procedure of validation cohort | ✓ ▪ |  |  |  |
| · Inclusion criteria | ✓ ▪ |  |  |  |
| · Exclusion criteria | ✓ ▪ |  |  |  |
| Describe patient sampling (random, consecutive, all, etc.) | ✓ ▪ |  |  |  |
| Describe data collection | ✓ ▪ |  |  |  |
| · Who identified patients and did selection adhere to patient recruitment criteria | ✓ ▪ |  |  |  |
| · Who collected data | ✓ ▪ |  |  |  |
| · A *priori* data collection form | ✓ ▪ |  |  |  |
| · Disease classification |  |  |  | ✓ ▪ |
| · Split sample (i.e. re-validation using a separate cohort) |  |  |  | ✓ ▪ |
| METHODS (cont.): |  |  |  |  |
| *Test Methods:* |  |  |  |  |
| Describe number, training and expertise of persons reading reference standard | ✓ ▪ |  |  |  |
| If >1 person reading reference standard, quote measure of consistency (e.g. kappa) | ✓ ▪ |  |  |  |
| Blinding of interpreters of reference standard to results of classification by administrative data e.g. Chart abstractor blinded to how that chart was coded |  |  | ✓ ▪ |  |
| *Statistical Methods:* |  |  |  |  |
| Describe methods of calculating/comparing diagnostic accuracy | ✓ |  |  |  |
| RESULTS: |  |  |  |  |
| *Participants:* |  |  |  |  |
| Report when study done, start/end dates of enrollment | ✓ |  |  |  |
| Describe number of people who satisfied inclusion/exclusion criteria | ✓ |  |  |  |
| Study flow diagram |  |  |  | ✓ |
| *Test results:* |  |  |  |  |
| Report distribution of disease severity |  |  |  | ✓ |
| Report cross-tabulation of index tests by results of reference standard | ✓ |  |  |  |
| RESULTS (cont.): |  |  |  |  |
| *Estimates:* |  |  |  |  |
| Report at least 4 estimates of diagnostic accuracy |  |  |  | ✓ |
| Diagnostic Accuracy Measures Reported: | ✓ |  |  |  |
| · Sensitivity |  |  |  | ✓ |
| · Spec |  |  |  | ✓ |
| · PPV | ✓ |  |  |  |
| · NPV |  |  |  | ✓ ▪ |
| · Likelihood ratios |  |  |  | ✓ ▪ |
| · Kappa | ✓ ▪ |  |  |  |
| · Area under the ROC curve / c-statistic |  |  |  | ✓ ▪ |
| · Accuracy/agreement | ✓ ▪ |  |  |  |
| · Other (specify) |  |  |  | ✓ ▪ |
| Report accuracy for subgroups (e.g. age, geography, different sex, etc.) |  |  |  | ✓ ▪ |
| If PPV/NPV reported, ratio of cases/controls of validation cohort approximate prevalence of condition in the population |  |  | ✓ ▪ |  |
| Report 95% confidence intervals for each diagnostic measure | ✓ ▪ |  |  |  |
| DISCUSSION: |  |  |  |  |
| Discuss the applicability of the validation findings | ✓ ▪ |  |  |  |

